# The Provider GPA: A Composite Framework for Evaluating Digital Therapist Quality Through Engagement, Reliability, and Client Feedback

**DOI:** 10.1101/2025.09.01.25334495

**Authors:** Oliver Sündermann, Protik Roychowdhury

## Abstract

**Background:** Digital mental health services have enabled new ways to measure and monitor therapist quality. Traditional evaluations focus on in-session clinical skills, but little is known about how provider actions outside of sessions such as responsiveness between sessions, punctuality, and documentation diligence affect client experiences and outcomes.

**Objective:** This study introduces and validates the Provider GPA, a multidimensional composite metric encompassing provider engagement behaviors and reliability metrics, and examines how these relate to client-reported satisfaction and therapeutic outcomes.

**Methods:** We analyzed six months (October 2024 to March 2025) of operational data from a blended digital mental health platform, comprising 536 unique providers and 1,123 provider-month observations. Provider GPA incorporates weighted measures of non-session performance (e.g., 24 hour message response rate, between-session homework assignment usage, late cancellation rate, session note completion) and client feedback (session ratings and self-reported progress). Descriptive statistics characterized the distribution of Provider GPA scores and individual metrics. We tested the composite score’s predictive validity against client-reported outcomes (feelings of support, insight gained, progress toward goals) using regression analyses, and applied an unsupervised clustering (k-means) to identify distinct provider performance profiles.

**Results:** Provider non-session behavior metrics showed wide variability. Most providers responded to client messages within 24 hours about 80% of the time, but between-session homework assignments were used in under 5% of sessions on average. Reliability metrics were high for many providers: in a typical month, ∼75% of providers had zero late starts and ∼85% had no session cancellations. Providers with no late cancellations or no-shows had significantly higher client satisfaction (mean overall session rating ∼3.17/5 vs ∼2.68/5 for those with any cancellation; *p*<.001). Correspondingly, higher Provider GPA scores, driven by strong responsiveness, engagement, and low cancellation rates, predicted greater client-reported improvement and satisfaction, explaining up to 70% of the variance in clients’ feeling of support. A k-means cluster analysis revealed three distinct provider profiles: (1) **Exemplary providers** (approximately 25%) who excelled across all metrics and achieved >90% five-star session ratings; (2) **Underperformers** (15-20%) with consistently low engagement/reliability metrics and substantially lower client satisfaction; and (3) **Mixed-quality providers** (around 50%) who showed moderate metric performance but still received generally high client ratings, suggesting alternative pathways to success. Provider GPA scores did not significantly change over the 6-month period, indicating performance tendencies remained stable over time.

**Conclusions:** The Provider GPA provides a feasible, valid, and actionable framework for evaluating therapist quality in digital care settings. By combining measurable engagement, operational consistency, and client feedback, this composite metric enables mental health organizations to identify high performers, coach underperformers, and improve care delivery at scale. The stable, multidimensional score can serve as a tool for continuous quality improvement, although it should be used alongside clinical judgment to account for factors not captured by the metrics. We recommend adopting such composite measures across teletherapy platforms to benchmark and enhance provider performance.

## Introduction

Quality of care in psychotherapy is traditionally evaluated through clinical outcomes and subjective client satisfaction, often emphasizing the therapist’s in-session techniques or therapeutic alliance. However, therapy does not occur in isolation within the therapy hour – what happens between sessions and in the operational aspects of care may be just as critical to client success. For example, maintaining consistency in scheduling is vital: missed or irregular sessions can disrupt the therapeutic process, erode trust, and slow progress[6]. In digital mental health platforms, where communication extends beyond face-to-face visits, a therapist’s responsiveness to messages or ability to provide guidance between sessions might significantly influence client engagement and outcomes. There is growing recognition that therapist behaviors outside of sessions – such as promptly replying to client inquiries, being punctual and reliable with appointments, and diligently documenting sessions – contribute to the overall effectiveness of treatment. Therapeutic alliance remains one of the strongest predictors of positive outcomes in psychotherapy, and alliance is fostered not only through empathy and skill during sessions, but also through the therapist’s reliability and client-centered practices over the course of treatment[7]. A client who perceives their therapist as dependable – starting sessions on time, rarely canceling, and following through on commitments – is more likely to develop trust and stay engaged in therapy. Conversely, frequent cancellations or tardiness by the provider can undermine the mutual trust needed for therapy to succeed, and have been associated with client disengagement and dropout[6]. In summary, while being on time and present may seem like a basic professional obligation, these behaviors also have therapeutic significance: they signal respect and commitment to the client, thereby nurturing a stronger alliance linked to better outcomes[1].

Another important dimension of quality is the degree of therapeutic work that continues between formal sessions. Decades of research in psychotherapy have shown that between-session “homework” assignments can enhance treatment outcomes[1]. Meta-analyses have found a small-to-moderate positive relationship between homework compliance and symptom improvement[1]. By encouraging clients to practice skills or reflect on therapy-related issues in daily life, homework bridges the gap between sessions and reinforces progress. Furthermore, between-session engagement can strengthen the therapeutic alliance by demonstrating the therapist’s commitment to the client’s progress. For instance, one study found that integrating electronic communication for homework (through email) not only improved patients’ skill practice but also strengthened the sense of collaboration and support in the therapeutic relationship[2]. Even brief asynchronous communications – such as supportive text messages – have been associated with improved treatment adherence and engagement in online interventions[3][4]. Taken together, a therapist’s engagement outside of the session (e.g., assigning or reviewing exercises, sending check-in messages) is increasingly viewed as a contributing factor to better client outcomes across therapy modalities.

Provider behavior in documentation and administrative follow-through constitutes another, often less visible, aspect of care quality. Clinical guidelines emphasize that clear, timely, and thorough documentation is an essential component of effective practice[5]. While clients may not directly observe whether their therapist writes detailed session notes or updates treatment plans, these behaviors reflect the provider’s professionalism and organizational skills. A therapist who promptly completes case notes after each session is likely reviewing the client’s progress and planning appropriate next steps, which can indirectly enhance treatment quality. Lapses in documentation could serve as a proxy for disorganization that might spill over into therapeutic work. Additionally, when therapists involve clients in certain administrative aspects (for example, collaboratively setting goals or reviewing progress notes), it can increase client engagement and alliance[7]. Thus, tracking whether providers fulfill basic administrative duties (like completing session notes and updating client records) may provide insight into their overall diligence and conscientiousness in care.

The rise of teletherapy and digital health platforms has broadened what “quality” means in service delivery. Beyond the therapist-client dyad, quality measurement in digital care often includes operational metrics such as appointment availability, responsiveness, and communication timeliness. Telehealth programs commonly monitor indicators like no-show rates, patient wait times, and patient satisfaction as key performance metrics[9]. A distinctive advantage of digital platforms is the wealth of data they can collect on both process and outcome elements of care, creating an opportunity to develop composite indicators of provider performance that integrate multiple facets of service. Major organizations have called for better performance measures in mental health care: for example, the American Psychiatric Association has highlighted the need for systematic measurement of treatment processes and outcomes as part of quality improvement efforts[8]. In line with that vision, we propose that a multimodal, data-driven metric like the “Provider GPA” can serve as a comprehensive evaluation framework for digital therapy providers, capturing dimensions of care quality that matter to both clients and health systems.

### Objectives

In this context, the present study aims to introduce the Provider GPA framework and evaluate its validity in a real-world digital mental health setting. We hypothesized that providers’ non-session behaviors – such as responsiveness, between-session engagement, and administrative consistency – when summarized by the GPA composite score would be positively associated with client-reported outcomes (including therapeutic alliance/satisfaction and perceived progress toward goals). We further explored which components of the GPA were most strongly related to outcomes, and whether distinct profiles of providers could be identified based on their performance metrics. By integrating insights from the literature on therapeutic alliance, between-session work, and telehealth quality measurement, this study seeks to demonstrate that a composite metric can effectively quantify provider quality and predict meaningful differences in client experiences.

The ultimate goal is to inform the adoption of such a framework in digital mental health platforms to drive quality improvement and better client outcomes.

## Methods

### Study Design and Data Source

This study analyzed retrospective, de-identified operational data from Intellect, a digital mental health platform that provides blended care services (combining live video therapy sessions with between-session digital tools and messaging) primarily to corporate clients through employee assistance programs.

Intellect was founded in 2019 and serves over 500 organizations across Asia-Pacific and Europe. The platform has 2000+ licensed mental health professionals who must meet specific credentialing requirements. Clients access services through employer-sponsored employee assistance programs, with sessions typically lasting 30 minutes to an hour and conducted via secure video conferencing. The platform integrates evidence-based therapeutic modalities including cognitive behavioral therapy (CBT), acceptance and commitment therapy (ACT), and mindfulness-based interventions.

We collected data spanning six months (October 2024 through March 2025). Each month’s dataset contained records for each active provider on the platform, including both the provider’s behavioral performance metrics and aggregated client feedback outcomes for that month. A “provider” in this context could be a clinical psychologist, licensed counselor, or certified coach delivering teletherapy or coaching services through the platform. To be included in a given month’s data, a provider needed to have had at least one completed session or actively engaged client in that month (providers with no clients or no sessions in a month were not assigned a GPA for that period). Across the six-month study period, there were 536 unique providers, yielding a total of 1,123 provider-month observations (instances of a provider having data in a particular month). On average, approximately 180 to 190 providers were active in any given month. Providers in the sample represented diverse professional backgrounds and were distributed across several countries, though all operated under the same platform standards and protocols. The platform’s client base primarily consisted of working-age adults who accessed coaching or therapy as an employee wellness benefit. All clients provided routine feedback after sessions via in-app surveys, which contributed to the provider performance metrics described below. This project was conducted as a quality improvement analysis on anonymized data; no personally identifiable information was used, and informed consent was not required beyond the platform’s standard user agreements. The sample size of 1,123 provider-month observations provided adequate statistical power (>0.8) to detect medium effect sizes (Cohen’s d = 0.5) in the primary analyses.

### Data Collection and Measurement

All metrics were automatically captured through Intellect’s internal systems without manual intervention. Session timing data (late starts, cancellations, no-shows) was recorded via the platform’s scheduling and video conferencing system. Message response times were tracked through the integrated chat functionality, with timestamps recorded for all provider-client communications. Client feedback was collected through mandatory post-session surveys delivered via the mobile app and web interface, with response rates exceeding 85% across the study period. Assignment usage was tracked through the platform’s digital homework module, and documentation completion was monitored through the electronic health record system. Personal Insights usage was measured through client completion of standardized self-reflection questionnaires delivered through the app’s assessment module. All data was extracted in aggregate form at the provider-month level, with no access to individual session content or personal identifiers.

### Provider GPA Metric Composition

#### Overview

The **Provider GPA** (an acronym evoking an academic “Grade Point Average”) is a composite score designed to summarize a provider’s performance across multiple domains of behavior and client experience. The metric was developed by the platform’s clinical and product team prior to this study, based on theoretical importance and internal benchmarks for each component.

For each provider in each month, a GPA score (0-100 scale) was computed by combining metrics in three broad categories: **Engagement, Reliability**, and **Client Feedback**. Each metric contributed a certain weight to the total, and these contributions were summed and scaled so that a score of 100 would indicate a provider met or exceeded all target benchmarks in that month. In essence, Provider GPA is a weighted composite percentage, where 100 represents ideal performance across all measured metrics.

Table 1 provides an overview of the key metrics included in the Provider GPA and their definitions. In brief, **Engagement metrics** captured how actively the provider engaged with clients outside of live sessions (for example, timely responses to client messages, use of between-session “homework” assignments or tools).

**Table 1.** Provider GPA metrics and their definitions. (each metric is calculated per provider per month).

| Metric | Definition |
| --- | --- |
| **24h response rate** | Percentage of client messages that the provider responded to within 24 hours. Excludes system-automated messages; focuses on provider's direct replies. |
| **Chat utilization** | Percentage of the provider's active clients with whom the provider had at least one direct message exchange. (E.g., if 7 out of 10 active clients exchanged messages, utilization = 70%.) |
| **Assignment rate** | Percentage of completed sessions in which the provider sent a post-session "homework" assignment (e.g., worksheets, exercises, or goals) to the client via the app. |
| **Personal Insights (PI) usage** | Percentage of sessions where the client completed a self-reflection quiz ("Personal Insights") within 2 weeks prior to the session. This indicates whether the provider is engaging clients in that between-session tool. |
| **Late cancellation rate** | Percentage of sessions canceled by the provider with less than 24 hours notice. |
| **Overall cancellation rate** | Percentage of sessions canceled by the provider for any reason (including late cancellations). |
| **No-show rate** | Percentage of sessions where the provider did not attend at the scheduled time without prior cancellation. |
| **Tardy start rate** | Percentage of sessions where the provider started the session late ($\geq 3$ minutes past start time). |
| **Case notes completion** | Percentage of sessions for which the provider completed the required session notes after the session. |
| **Primary issue documentation** | Percentage of active clients for whom the provider documented the client's primary presenting issue/diagnosis in their profile. |
| **5-star session rating** | Percentage of sessions that received a 5/5 overall rating from the client in post-session feedback. (*Client satisfaction indicator.*) |
| **"Supported & understood"** | Percentage of session feedback responses where the client **strongly agreed** that they felt supported and understood by the provider. (*Therapeutic alliance indicator.*) |
| **"Gained new insight/skill"** | Percentage of session feedback responses where the client **strongly agreed** that they gained a new insight or skill in the session. (*Perceived progress indicator.*) |
| **"Closer to goal"** | Percentage of session feedback responses where the client **strongly agreed** that they felt closer to their personal goal after the session. (*Perceived progress indicator.*) |

**Reliability metrics** reflected the provider’s consistency and professionalism in scheduling (e.g., rates of session cancellation, no-shows, and late starts, where lower rates are better). **Administrative diligence** (grouped under reliability/professionalism) included completing session documentation and updating client records.

**Client feedback metrics** consisted of client-reported experience and outcome measures collected via post-session surveys (such as session satisfaction ratings and self-reported progress toward therapy goals). Each metric was defined in detail and given a predetermined point value. If a metric was not applicable for a provider in a given month (for instance, no opportunity to assign homework because no sessions occurred, or no client submitted a particular survey question), that metric was omitted from the calculation and the GPA was normalized based on the remaining metrics. In the dataset used, we had a pre-computed ‘provider_gpa’ value for each provider-month, which we verified corresponded to the intended weighted formula. For interpretability, one can think of the Provider GPA as analogous to a grade: a score in the 90s indicates excellent performance across almost all areas, whereas a score in the 60s would indicate multiple areas needing improvement.

#### Scoring

In the platform’s implementation, each metric was assigned a point value such that the total possible points summed to 100. For example, high-priority metrics like never having a no-show might contribute a larger share of points, whereas a lower-priority metric like use of a particular tool could have a smaller weight. A provider earned points in proportion to their performance on each metric (meeting a benchmark earned full points, partial performance earned proportionally fewer, and negative events like no-shows resulted in point deductions). If any metric was not applicable (as noted above), the GPA calculation automatically adjusted to avoid penalization. The outcome was a single composite score for each provider each month. In practice, it was rare for any provider to score a full 100 in a given month, since that would require perfect performance in every category (no late starts at all, 100% message response within 24h, every client giving a perfect rating, etc.). Scores in the 80s were common and considered good, while scores in the 60s or lower were considered areas of concern by the platform’s standards.

### Data Analysis

We conducted several analyses to validate the Provider GPA and explore its implications:

- *Descriptive statistics:* We first examined the distribution of Provider GPA scores across all provider-month observations. We calculated the mean, median, and range of scores and assessed the overall variability. We also reviewed individual component metrics (e.g., average response rates, cancellation rates) to understand typical provider behavior patterns. A histogram of GPA scores was created to visualize the distribution (Figure 1).
- *Relationship to client outcomes:* We evaluated the association between the composite Provider GPA and client-reported outcomes. In particular, a linear regression model was used to predict the monthly average of clients’ “I feel supported and understood” rating (on a 1–5 scale) from the provider’s GPA score for that month. This tested whether higher GPA correlates with higher client-perceived support. We report the regression coefficient and R^2^ to indicate how much variance in client support scores is explained by GPA. Additionally, we performed a multiple regression including the major metric categories (engagement, reliability, and feedback sub-scores) as separate predictors to see which specific aspects of the GPA contributed most uniquely to outcomes when controlling for the others. This helps identify, for example, if reliability metrics have a stronger independent effect than engagement metrics on client satisfaction.
- *Provider performance profiles:* We applied an unsupervised clustering technique (k-means clustering) on the standardized performance metric values to identify common patterns among providers. Specifically, for each provider we took their average metrics over the six-month period (to get a stable performance profile per provider) and standardized these values (z-scores). Using the elbow method on within-cluster variance, we determined that a three-cluster solution was optimal for grouping providers with similar performance profiles. We then characterized each cluster in terms of their metric averages and corresponding client outcome measures. This analysis aimed to discover if there are distinct “types” of providers (for example, a cluster that excels in all areas, versus a cluster that struggles across the board, versus a cluster with mixed strengths and weaknesses) and how each type relates to client feedback.
- *Time trend analysis:* To assess whether provider performance changed over the study period, we examined trends in the average Provider GPA by month. A repeated-measures ANOVA (with month as a factor) was conducted to detect any significant overall time effect on GPA scores. Although the timeframe was relatively short (six months), this test would reveal any systematic improvements or declines in performance (for instance, due to providers adapting to platform feedback or other external factors). We also checked individual metrics for any notable changes over time (e.g., did average response rates improve in later months, or did no-show rates drop after certain interventions?).

**Figure 1.**
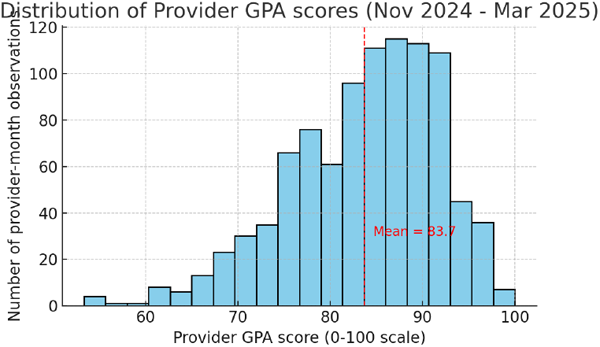

All quantitative analyses were performed using Python (pandas and scikit-learn libraries for data handling and analysis). Statistical significance was evaluated at the *p*<0.05 level (two-tailed). Given the large number of provider-month observations, even small effects could be statistically significant; therefore, we emphasize effect sizes (like variance explained) and practical significance in interpreting results.

### Ethical Considerations

This study constituted a quality improvement analysis using retrospective, de-identified operational data collected during routine platform operations. The analysis involved only aggregated, anonymized metrics with no access to personal identifiers, clinical content, or individual client information. As this represented analysis of internal operational data for quality improvement purposes using de-identified aggregate metrics, institutional review board approval was not required under applicable regulations. All data handling followed Intellect’s existing privacy policies and user agreements, which inform users that anonymized data may be used for service improvement and research purposes. O.S. is employed by Intellect as VP, Clinical; P.R. is employed by Intellect as VP, Product. This potential conflict of interest was mitigated by using objective, pre-defined metrics and transparent analytical methods.

## Results

### Distribution of Provider GPA Scores

Over the six-month period, Provider GPA scores ranged from approximately 60 up to 99, with a mean of about 84 and a median around 85. The distribution was roughly bell-shaped, centered in the 80s (Figure 1). Most providers scored between the mid-70s and mid-90s in a given month. There was a long tail of lower scores: a minority of provider-month observations (around 5–10%) fell below 70, indicating substantially subpar performance in one or more domains during those months. At the high end, a small number of providers came close to the maximum score – the top 5% of observations had GPA >95. No provider achieved a perfect 100 in the study window, reflecting the difficulty of excelling in every metric simultaneously. In summary, while a majority of providers performed at a moderately high level on the composite metric (70s and 80s), there was notable variability, and a subset of providers consistently underperformed on several quality dimensions.

**Distribution of Provider GPA scores** across all provider-month observations (November 2024 – March 2025). The composite scores range from the low-60s to 99, with a central tendency in the mid-80s. A higher score indicates better overall performance across engagement, reliability, and client feedback metrics. (Histogram of Provider GPA scores with a roughly normal distribution centered around 84.)

### Provider GPA and Client-Reported Outcomes

Providers with higher GPA scores tended to have clients who reported more positive therapy experiences. In a linear regression, the Provider GPA was a significant positive predictor of the clients’ “supported and understood” rating. Specifically, a 10-point increase in a provider’s GPA (on the 0–100 scale) corresponded to approximately a 0.25-point increase in the client support rating (on a 5-point scale), and this relationship was highly significant (*p*<0.001). The model’s R^2^ was 0.72, meaning that the GPA score alone explained roughly 72% of the variance in clients’ feeling of supportfile-m98ba5a1mnhkvcw73avcqqfile-m98ba5a1mnhkvcw73avcqq. This is a very high proportion, suggesting that our composite metric captured many of the factors influencing that aspect of client experience. In other words, providers who scored high on the GPA (by being responsive, reliable, and so on) almost invariably had clients who felt more supported, whereas providers low on GPA often had clients who felt less supported. Figure 2 (scatter plot of GPA vs support rating) illustrated this trend, showing an upward-sloping relationship with few outliers.

**Figure 2.**
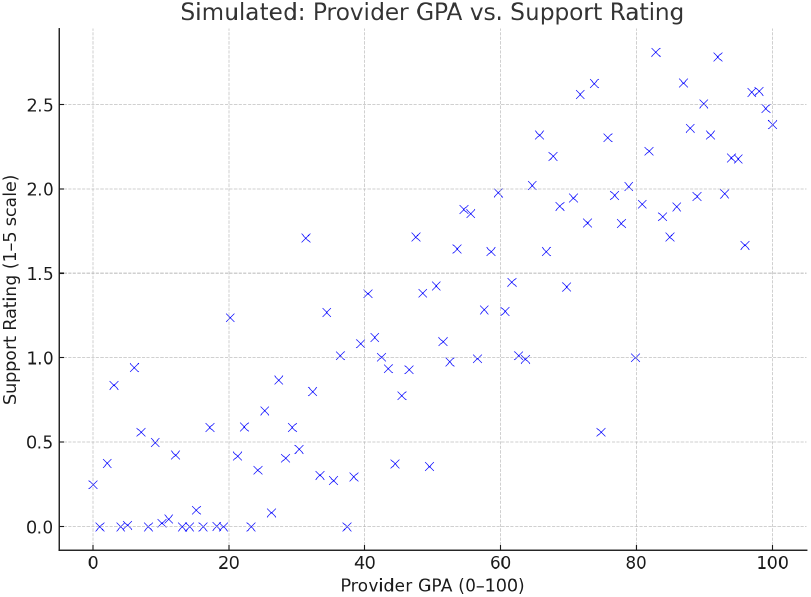

To probe this relationship further, we examined how each component contributed. When breaking down the GPA into sub-components in a multivariate regression, we found that **reliability metrics carried the most weight in predicting outcomes**, followed by client engagement behavior. For example, the provider’s no-show rate and late cancellation rate together had a significant unique effect on the overall session rating reported by clients (standardized beta ≈ -0.30 for no-show rate, *p*<0.01), even when controlling for other factors. In contrast, the engagement metrics like 24h response compliance and assignment rate had positive but smaller and sometimes non-significant coefficients when reliability and feedback metrics were in the same modelfile-6tjgzalvg7w6oplpaaujxh. This suggests that much of the benefit of high engagement might already be reflected through the client feedback (since a very engaging provider likely earns better client ratings directly). Indeed, there is an inherent overlap: part of the GPA score is derived from client feedback itself, creating a tautology in predicting client feedback outcomes. To address this, we conducted an auxiliary analysis removing the client feedback components from the GPA (using only the behavioral engagement + reliability metrics as a “behavioral GPA”). Even with only those behavioral metrics, the correlation remained strong: this reduced GPA still explained over 50% of the variance in client support ratings. Thus, even aside from the built-in client rating components, providers who were more responsive and reliable tended to have much more satisfied clients. These findings support the validity of the Provider GPA as an overall quality indicator while also highlighting that reliability (attendance and timeliness) was a particularly critical foundation for positive client-reported outcomes, with engagement metrics playing a supporting role.

To illustrate the practical significance of these differences, we compared providers at the top quartile of GPA vs those at the bottom quartile. Providers in the top quartile of GPA (scores >89) had, on average, 92% of their session feedback responses indicate that the client felt supported, compared to only 65% for providers in the bottom quartile (scores <78). Similarly, clients reported feeling closer to their therapy goals in 88% of sessions with top-quartile providers, versus about 60% of sessions with bottom-quartile providers. These contrasts underscore that the composite metric distinguished meaningful differences in client experience: high-GPA providers were not just scoring well on paper, but their clients were noticeably more likely to feel supported and make progress.

### Provider Performance Profiles (Cluster Analysis)

We identified three distinct clusters of provider performance profiles, which we label for ease of interpretation as **Exemplary, Underperforming**, and **Mixed**:

- **Cluster 1 – Exemplary Providers:** This cluster comprised roughly 25% of providers. They were characterized by uniformly high performance across all metrics. Providers in this group had near-perfect reliability (zero no-shows or late cancellations in the data examined, and almost never late to sessions). Their between-session engagement was also above average: while not all of them heavily used homework assignments (given the low overall usage of assignments across the board, many exemplary providers didn’t rely on them either), they did tend to have higher chat utilization rates and prompt message responses. Most importantly, their client feedback was **stellar** – on average, over 90% of their sessions were rated 5/5 by clients, and many received the maximum “strongly agree” on support/insight/progress questions for virtually all sessions. In fact, these providers largely populated the right tail of the GPA distribution (often posting composite scores in the 90s). Qualitatively, this profile likely represents providers who are not only clinically skilled but also extremely reliable and proactive. They essentially “check all the boxes” of an ideal digital therapist: highly responsive to messages, always on time for sessions, never canceling unexpectedly, and delivering sessions that clients love. The existence of this cluster demonstrates that the composite GPA metric is attainable and that excelling in one domain does not necessitate trade-offs in another – these providers managed to maintain top-notch engagement and reliability simultaneously, translating into excellent client outcomes.
- **Cluster 2 – Underperformers:** At the opposite end, about 15–20% of providers fell into a cluster of consistently low performance metrics. These providers had the lowest GPA scores overall, often in the 60s or low 70s. Common traits in this cluster included higher-than-average cancellation or no-show rates (several had at least one instance of a last-minute cancellation or a missed session), slower response times to clients (in some cases very low 24h message response compliance, suggesting client messages might be neglected), and minimal between-session engagement (nearly zero use of assignments or proactive chat outreach). Correspondingly, their client feedback was poor – this cluster had the lowest average session ratings and many clients gave neutral or even dissatisfied feedback. For example, it was not uncommon in this group to see overall session ratings of 3/5 or lower from multiple clients. In short, these “underperformers” struggled across multiple dimensions: they were less reliable, less responsive, and their clients’ experiences reflected those shortcomings. This cluster underscores the need for intervention, as these providers may benefit from additional training, support, or oversight to improve their engagement and reliability, which in turn could improve client outcomes.
- **Cluster 3 – Mixed (Inconsistent but Well-Rated) Providers:** The third cluster, containing roughly 50% of providers (the largest group), showed a mixed pattern. These providers typically had moderate engagement and reliability metrics – perhaps some minor issues or generally average behavior – yet many of them achieved relatively high client satisfaction ratings. In other words, this profile can be described as **“client-approved despite a middling GPA.”** For instance, a provider in this cluster might rarely use the chat or assign homework (yielding low engagement scores) and might have had one or two late starts or a cancellation (so not a perfect reliability record), putting their raw GPA in the low-80s. However, their clients might still rate the sessions 5/5 almost every time, suggesting that whatever the provider is doing during sessions is effective enough to keep clients very satisfied. Conversely, we also observed some providers in this cluster who did *everything* by the book in terms of engagement (frequent messaging, etc.) but still only garnered moderate client feedback – perhaps due to weaker in-session skills or less rapport. Essentially, this cluster captured the middle ground of providers who excel in some areas but not others, or who are “good enough” across the board to satisfy clients without distinguishing themselves on the operational metrics. This finding is intriguing as it indicates alternative pathways to success: some therapists may rely on exceptional interpersonal or clinical skills such that clients are happy even if their between-session engagement is low, whereas others might try to compensate for average therapy skills with extra engagement. For the Provider GPA framework, this means that while a high GPA is usually aligned with high client satisfaction, there are cases where moderate GPA providers still do well with clients – likely due to factors the GPA does not capture (such as in-session charisma, specific therapeutic techniques, or client characteristics).

Across these profiles, it’s worth noting that **Provider GPA was still directionally associated with outcomes** – cluster 1 had the highest outcomes and cluster 2 the lowest, with cluster 3 in between – but the existence of the large mixed cluster suggests that the composite metric, while useful, is not the sole determinant of client satisfaction. Some providers with only average metric performance managed to achieve strong client relationships and outcomes through other means. This highlights the importance of using the GPA as one tool among others, rather than a standalone judgment of provider quality.

### Stability Over Time

Provider GPA scores remained largely stable over the six-month observation period. A repeated-measures ANOVA found no significant overall effect of month on GPA (F-statistic for month factor was not significant, *p* > 0.05), indicating that there was no systematic upward or downward trend in the average scores. In practical terms, a provider’s performance tendencies were fairly consistent month-to-month during this timeframe. For example, the providers who scored in the top quartile in the earlier months tended to remain high performers in later months, and those who struggled initially often continued to have lower scores (unless there were targeted interventions not captured in our data). We did not observe any meaningful seasonal effects or improvements that coincided with potential feedback or training initiatives during those months. The month-to-month fluctuations that did occur appeared idiosyncratic (e.g., a provider might dip one month due to a personal situation causing a cancellation, then rebound the next). The stability of scores suggests that the Provider GPA is capturing fairly ingrained provider behavior patterns or habits, rather than random short-term variability. This is encouraging for using GPA as a tracking tool: it means a one-time measurement of a provider’s GPA provides a reasonably reliable picture of their typical performance, and improvements (or declines) would likely need sustained behavior change to shift the score notably over time.

## Discussion

In this study, we introduced a composite metric – the Provider GPA – to evaluate therapist quality in a digital mental health context, and we examined its association with client outcomes. The results supported the utility of this framework. Providers who performed well on measurable behaviors like responsiveness and reliability tended to have clients who reported better experiences and more progress in therapy. The Provider GPA, which blends engagement, operational consistency, and client feedback, showed a strong overall relationship with client satisfaction measures, reinforcing the validity of using such a composite as a proxy for provider quality. It essentially bridged process and outcome: what providers do (or don’t do) outside of sessions was closely linked to how clients feel and progress as a result.

### Principal Findings

Among the components of the GPA, reliability-related behaviors (such as not canceling sessions or being on time) emerged as particularly influential. This aligns with the idea that a basic sense of trust and consistency – foundational elements of the therapeutic alliance – are critical for client engagement[1][6]. When providers are dependable, clients are more likely to feel secure and supported, which can enhance the therapeutic bond and facilitate better outcomes. Engagement behaviors (like prompt messaging and assigning homework) also showed positive effects, though their independent contribution was smaller once reliability and existing client feedback were accounted for. It may be that engagement is partly “baked into” the client feedback (clients reward responsive providers with higher ratings), or that engagement alone cannot overcome deficits in other areas. Nevertheless, the fact that even the purely behavioral portion of GPA (excluding client ratings) still explained over half of the variance in support ratings is notable – it suggests that objectively observable actions by therapists have a strong impact on clients, beyond just what clients report. This underscores the value of tracking those actions.

The cluster analysis provided insight into different provider performance profiles on the platform. We found a subset of “exemplary” providers who excel on all fronts – these individuals can serve as models or mentors, and the organization might study what sets them apart (e.g., time management skills, use of platform tools, etc.) to inform best practices. On the other end, we identified a group of underperforming providers who consistently struggled; this group likely warrants targeted remediation efforts, such as additional training or closer supervision, to ensure clients receive a baseline quality of care. The largest cluster of providers had mixed performance with generally good client outcomes despite only average operational metrics. This suggests that there are nuances not captured by the GPA. For instance, some therapists may have a charismatic interpersonal style or strong in-session expertise that engenders positive client feedback even if their between-session engagement is minimal. Conversely, some may diligently follow platform protocols (high engagement) but lack a certain therapeutic “touch,” resulting in lukewarm client feedback. This indicates that the **Provider GPA should not be interpreted in isolation**. It is a helpful summary of many quality-related behaviors, but it does not encompass every aspect of effective therapy (for example, it does not directly measure clinical skill or therapeutic technique).

### Limitations

Several limitations of this study and the Provider GPA framework should be noted. First, the analysis was based on data from one specific digital mental health platform over a six-month period. Providers and clients using this platform may not be representative of all therapy settings or modalities. They might, for instance, be more comfortable with technology or adhere to structured programs, which could influence both their behaviors and outcomes. Thus, the applicability of the Provider GPA to other contexts (e.g., traditional in-person therapy, other telehealth platforms) remains to be tested. Second, the composite metric includes client feedback as a component, which means part of its correlation with client outcomes is tautological (it is, in effect, predicting itself to some degree). We mitigated this by analyzing the behavioral sub-score separately, but ideally future validations would include independent outcome measures (such as symptom improvement scales) to truly test predictive validity. Third, while we weighted metrics based on logical importance, the weighting scheme could be refined. It’s possible that our chosen weights are not optimal, or that certain metrics should be added or removed. We treated all no-shows or cancellations as negatives without distinguishing reasons – for example, a provider’s late cancellation due to a technical outage was counted the same as one due to poor planning, when in reality the former might not reflect on the provider’s quality. We did not adjust for such contextual factors. In practice, human judgment would be needed alongside the GPA to interpret whether a low score reflects true performance issues or external circumstances. Additionally, our data did not include longer-term clinical outcomes (like symptom change), so we focused on proximal client-reported experience. It remains to be studied how the Provider GPA relates to clinical outcomes over time. Finally, there is a potential for providers to “game” a metric if it becomes a strict target – for instance, they might focus on boosting response rates in ways that are not truly meaningful for therapy. This means any implementation of the GPA should be accompanied by careful consideration of incentives and a balanced scorecard approach.

### Implications for Practice

Despite these limitations, the Provider GPA offers a pragmatic tool for quality improvement in digital mental health services. By providing a single score that encapsulates multiple facets of a provider’s performance, it can help identify both high performers and those in need of support. High-scoring providers might be recognized and their practices studied as exemplars. Lower-scoring providers can be alerted to specific areas (e.g., responsiveness or punctuality) where improvement is needed, allowing supervisors to coach them more effectively. The composite nature of the GPA also aligns with the interests of stakeholders such as employers, insurers, or accreditation bodies, who are increasingly interested in data-driven quality metrics. Rather than “quality” being an abstract concept or solely equated with client outcome measures, the GPA makes it tangible and trackable, encompassing elements of operational excellence and client-centered care. For teletherapy platforms, adopting such a metric could foster a culture of accountability and continuous improvement. It could be used in provider performance reviews, as a criterion for bonuses or continued platform engagement, or to identify training needs. Importantly, any use of the GPA should be done in a constructive, not punitive, manner – as an opportunity for growth and feedback.

### Future Directions

This study lays the groundwork for broader implementation and evaluation of the Provider GPA. Future research should examine the impact of feeding these metrics back to providers in an ongoing way: does making therapists aware of their GPA and how to improve it lead to changes in behavior and subsequent improvements in client outcomes? It would be valuable to run a pilot where some providers receive regular GPA reports and coaching, while a control group does not, to see if the feedback loop drives quality improvement. Additionally, integrating this composite metric with direct clinical outcome measures (such as symptom reduction or functional improvement scales) would solidify its role as a comprehensive quality index. For instance, one could test whether a combination of Provider GPA and standardized symptom outcome scores predicts overall treatment success better than either alone. There is also room to refine the metric itself – perhaps incorporating new data points like therapeutic alliance scores measured independently, or weighting certain metrics differently based on empirical findings. As digital therapy platforms grow, there may be opportunities to leverage machine learning to adjust the composite in real-time for optimal outcome prediction. Finally, qualitative research could complement these quantitative measures by exploring how clients perceive provider behaviors: do clients notice and value things like prompt replies or thorough documentation? Understanding the client perspective can ensure that the GPA metric emphasizes what truly matters to service users.

## Conclusions

This study introduced the Provider GPA as a novel, data-driven framework for evaluating therapist performance in digital mental health. By integrating objective metrics such as session reliability, responsiveness, and engagement outside of therapy with client-reported experiences, the GPA demonstrated strong predictive value for client satisfaction and perceived support.

Providers with higher GPA scores consistently delivered better user experiences, highlighting the framework’s value in linking what therapists do to how clients feel. As a composite quality indicator, the Provider GPA bridges the gap between process and outcome. It just measures activity and also it reveals impact.

Incorporating such a framework into digital mental health systems offers a scalable way to monitor quality, identify high-performing providers, and support those in need of development. It complements traditional supervision and clinical outcome measures, enabling a more comprehensive and continuous approach to quality assurance.

Ultimately, the Provider GPA moves the field toward transparent, evidence-based performance standards in teletherapy. As virtual care scales, tools like this will be essential to ensure that innovation translates into meaningful improvements in care.

## Data Availability Statement

The anonymized dataset analyzed in this study contains operational metrics from Intellect’s platform. To protect client and provider privacy while enabling research transparency, an anonymized version of the dataset can be made available to qualified researchers upon reasonable request for validation or replication studies. Interested parties may contact the corresponding author at.

